# Disease Characteristics Influence the Privacy Calculus to adopt Electronic Health Records: A randomized controlled trial in Germany

**DOI:** 10.1101/2024.02.07.24302380

**Authors:** Niklas von Kalckreuth, Markus Feufel

**Affiliations:** Department of Psychology and Ergonomics (IPA), Technische Universität Berlin, Straße des 17. Juni 135, 10623 Berlin, Germany

**Author notes:** Correspondence: Department of Psychology and Ergonomics (IPA), Technische Universität Berlin, Straße des 17. Juni 135, 10623 Berlin, Germany, ORCID: 0000-0001-9165-682X, ORCID: 0000-0003-0563-8831.

**Keywords:** mHealth, mobile health, privacy calculus, electronic health record, EHR, intention, disease characteristics, time course, adoption, benefit, privacy concern, trust, social norms, data autonomy, attitude to privacy

## Abstract

**Background:** The electronic health record (EHR) is integral to improving healthcare efficiency and quality. Its successful implementation hinges on patient willingness to use it, particularly in Germany where concerns about data security and privacy significantly influence usage intention. Little is known, however, about how specific characteristics of medical data influence patients’ intention to use the EHR.

**Objective:** This study aims to validate the Privacy Calculus Model (PCM) in the EHR context and to assess how personal and disease characteristics, namely disease-related stigma and disease time course, affect PCM predictions.

**Methods:** An online survey was conducted to empirically validate the PCM for EHR, incorporating a case vignette varying in disease-related stigma (high/low) and time course (acute/chronic), with 241 German participants. The data were analyzed using SEM-PLS.

**Results:** The model explains R²=71.8% of the variance in intention to use. The intention to use is influenced by perceived benefits, data privacy concerns, trust in the provider, and social norms. However, only the disease’s time course, not stigma, affects this intention. For acute diseases, perceived benefits and social norms are influential, whereas for chronic diseases, perceived benefits, privacy concerns, and trust in the provider influence intention.

**Conclusions:** The PCM validation for EHRs reveals that personal and disease characteristics shape usage intention in Germany. This suggests the need for tailored EHR adoption strategies that address specific needs and concerns of patients with different disease types. Such strategies could lead to a more successful and widespread implementation of EHRs, especially in privacy-conscious contexts.

## Introduction

### Background

Information technology used to collect, analyze and interpret health data is often seen as a solution for improving the efficiency and quality of healthcare delivery.^1,2^ An important facet of this digital transformation process is the electronic health record (EHR), in which patients’ health data (e.g. diagnoses, therapies, vaccinations, discharge reports, emergency data and medication plans^3,4^) can be documented, exchanged and viewed.^3,5^ As technology, the EHR promises to facilitate communication and coordination between healthcare professionals (e.g., physicians, therapists, pharmacists) and patients to, ultimately, improve patient care and reduce costs at the same time.^2,6^ For instance, pre-existing conditions, intolerances and medication plans could be considered during diagnosis and treatment to avoid double diagnoses, over-, and undertreatment.^5^ At the same time, an improved documentation may free time, which physicians could spend interacting with their patients.^7^

Although the EHR has been successfully implemented in countries such as Denmark, Finland and Estonia,^8,9^ implementation has been less successful in other countries (e.g. France, Australia) or is still in the implementation stage (e.g. in Germany). In Australia, for instance, 23 million EHRs have been created but remain empty and in France less than 9.5% of the population have created a record; even fewer have filled it with their health data.^10,11^ In Germany, 3 out of 4 citizens state that they would use the EHR^12^, but concerns about data security and data privacy of the EHR remain high.^7^ Ultimately, the success of the EHR seems to depend on whether and under what circumstances it is used by patients, all the more because it is the patient who decides what data are stored, shared and displayed in the EHR (cf. Patient Data Protection Act in Germany).^5,13^

Consequently, it is important to understand both which disease-specific *and* user-specific factors influence users’ intention to (not) use the EHR in order to improve the EHR and increase the number of users so that the EHR can realize its touted potential. In previous studies, we and others (a) have adapted the privacy calculus model to understand privacy concerns and intentions to use a technology in the healthcare domain^10,14,15^ and (b) have shown that the decisions to upload a medical report in the EHR are influenced by certain characteristics of the health data to be stored in the EHR, such as the time course and potential for stigmatization associated with the disease mentioned on a medical report.^7,16–18^ But and whether and how disease and user-specific characteristics influence the privacy calculus and the intention to use the EHR remains an open question.

### Prior Work

The privacy calculus model is an established model for examining intentions to use a technology when personal data are involved.^19,20^ The model, originally introduced to study interactions with social network sites (SNSs), assumes that users compare potential benefits and costs of using the technology. If the sum of the benefits exceeds the costs, people will use the technology. Vice versa, people will refuse to use it if costs outweigh benefits.^15,21,22^ In a preliminary study, we validated a privacy calculus model adapted for the healthcare domain with a health app of a health insurance.^14,23^ The adapted model suggests that patients’ intention to use mobile health (mHealth) apps is influenced not only by common privacy calculus factors^10,15^ such as perceived benefits, privacy concerns and trust in the provider, but also patients’ general attitudes to privacy, their sense of control over personal data and the societal acceptance of using these apps (social norms).

However, the intention to use mHealth applications is highly context-dependent.^24^ Although the two systems - EHR and health insurance apps - are rather comparable (users upload and manage sensitive health data in both systems), an important difference is that the health insurance app serves as a communication platform between patients and health insurance companies, whereas the EHR connects and shares patient data with *all* stakeholders in the healthcare system. This difference may result in difference privacy concerns. Thus, our goal for this article is first to validate the model, which was developed for mHealth technologies in general, for the specific EHR context.

Moreover, both user and disease characteristics tend to influence the decision to use mHealth and internet of things (IoT) technology.^25,26^ In previous studies, we have shown that also EHR usage depends on disease characteristics as well as on different use cases (e.g. multimorbid versus previously healthy patients).^16,17^ Whereas for multimorbid patients suffering from various acute and chronic diseases both the time course and the stigmatization potential of a disease had an influence on the decision to upload medical reports to the EHR,^16^ previously healthy patients changed their uploading behavior only for diseases with stigmatization potential.^17^ Hence, in this article, we examine the influence of both factors on the intention to use the EHR to help better understand user interaction with this central digital health technology.

### Aim of this Research and Approach

To explain the intention to use the EHR in terms of time course and stigmatization potential of diseases, we briefly review the existing privacy calculus model for the eHealth context and its hypotheses. We then validate the model for the EHR use case using structural equation modeling with partial least squares (SEM-PLS). We test the influence of disease and personal characteristics on the intention to use an EHR as predicted by the model using multigroup analysis (MGA). After discussing the results, we derive theoretical and practical implications and reflect on the limitations of the study. We end our paper with a conclusion.

### Model Description and Hypotheses

Figure 1 shows the research model of our previous study.^14,23^ As in the model already validated for mHealth applications, we assume that our outcome variable, intention to use the EHR, is influenced by perceived benefits, privacy concerns, trust in the provider, social norms (what we believe others think we should do as well as their actual behavior), attitude to privacy and perceived control over personal data. The derived hypotheses are summarized in Table 1.

**Figure 1.**
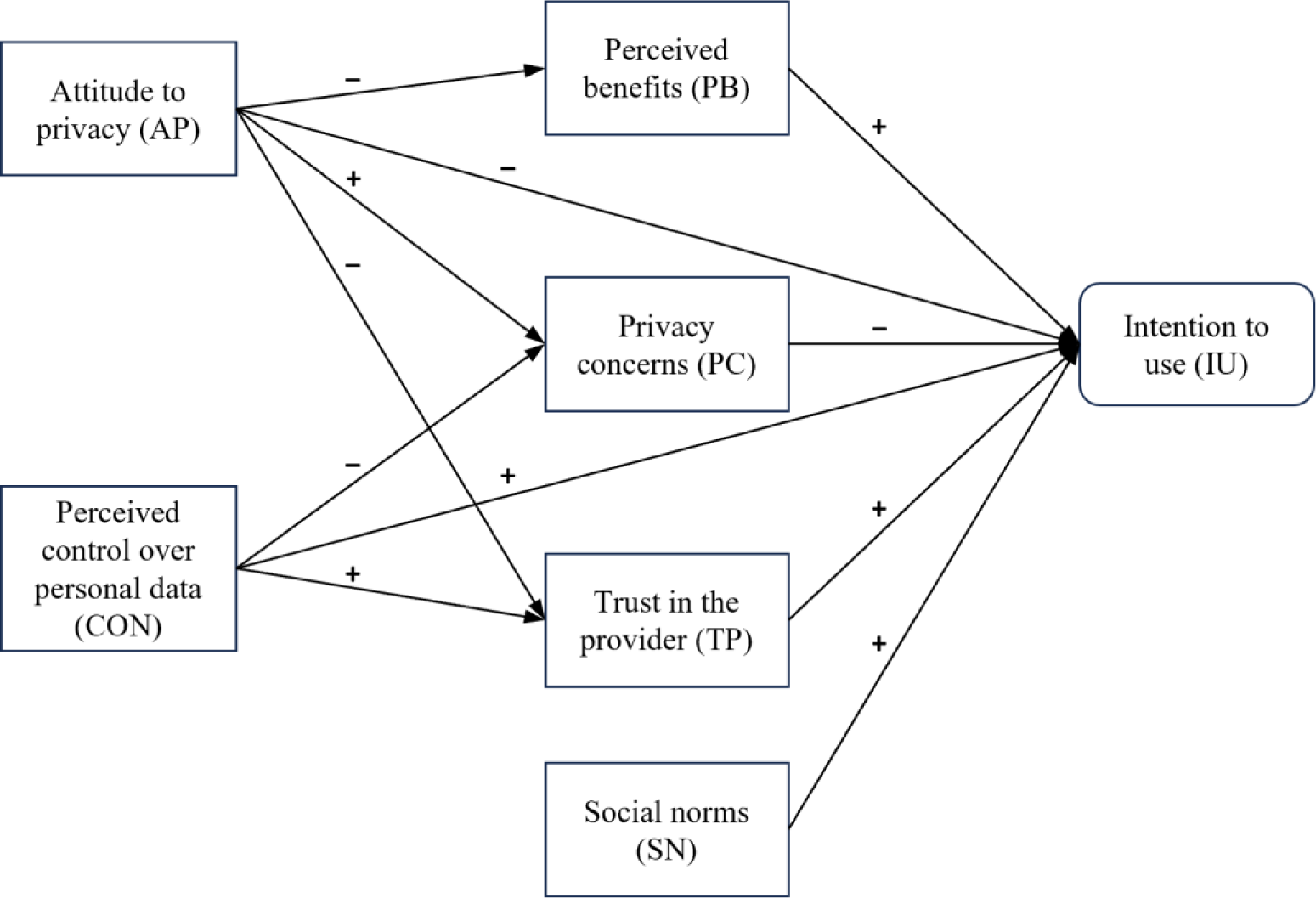
The privacy calculus model to predict intention to use mHealth apps^14^.

**Table 1.** Hypotheses regarding the research model.

| H# | Hypothesis |
| --- | --- |
| H1 | Perceived benefits positively influence users' intention to use the EHR. |
| H2 | Privacy concerns negatively influence users' intention to use the EHR. |
| H3 | Trust in the provider positively influences users' intention to use the EHR. |
| H4 | Social norms positively influence users' intention to use the EHR. |
| H5a | Perceived control over personal data positively influences users' intention to use the EHR. |
| H5b | Perceived control over personal data negatively influences users' privacy concerns. |
| H5c | Perceived control over personal data positively influences users' trust in the provider. |
| H6a | Attitude to privacy negatively influences users' intention to use the EHR. |
| H6b | Attitude to privacy negatively influences users' perceived benefits. |
| H6c | Attitude to privacy positively influences users' privacy concerns. |
| H6d | Attitude to privacy negatively influences users' trust in the provider. |

## Methods

### Ethics Approval and Consent to Participate

This study was approved by the Ethics Committee of the Department of Psychology and Ergonomics (Institut für Psychologie und Arbeitswissenschaft [IPA]) at Technische Universität Berlin (tracking number: AWB_KAL_1_230311). Participants volunteered to participate in the survey, and informed consent was required. On the first page of the survey, participants were told about the experimenter, the study purpose, what data were to be collected during the study, and where and for how long they would be stored. Hence, participants were informed about the duration of the survey (approximately 8 minutes) as well as the compensation for participation. Also, the participants had the possibility to download a pdf of the participant information on the first page.

### Participants

The model described in Figure 1 was empirically tested using data gathered via an online survey, that was performed as part of a bigger study, which compared the constructs intention to use and actual upload behavior. Following the “10 times rule”,^27^ we aimed for a total sample size of n=240 participants (n=60 participants for each of the 4 planned experimental conditions).

Individuals aged 18 years and older residing in Germany were allowed to participate in the study, as the content and questions of the study were designed to fit the context of the German EHR. Another prerequisite was that the participants had no personal previous experience (own illness) with the diseases mentioned in the respective medical reports, as the handling of stigmatized diseases by affected persons is different from that of unaffected persons.^28^ Sampling was done through Prolific, a clickworker platform characterized by high data quality.^29^ The study was conducted from May 9, 2023, till May 10, 2023. Participation was compensated with 1.60€, which corresponds to the German minimum wage. The mean value of the processing time was 9:28 minutes (SD 3:47 minutes) and the median 8:36 minutes. 275 individuals participated in the study.

### Design

We used a 2×2 between subject study design with the two independent variables (IVs) stigmatization potential and time course of the disease stated on a medical report. Stigmatization potential is associated with high risks^16,30^ that consequences could arise related to areas of personal lifestyle, occupation, and social life if medical findings became known.^16,28^ The time course is a classification of diseases in terms of their duration. These can be either *acute* (diseases of short duration that come on quickly) or *chronic* (slowly developing or long-lasting disease). In a preliminary study, respondents in a qualitative semi-structured interview described uploading medical findings of chronic diseases into the EHR as more beneficial than uploading findings of acute diseases.^16^ Participants were randomly assigned to one of these conditions (simple randomization, ratio: 1:1:1:1) using LimeSurvey’s built-in “rand” function. The dependent variables were the constructs of the privacy calculus model: intention to use, perceived benefits, privacy concerns, trust in the provider, social norms, attitude to privacy and perceived control over personal data.

### Materials

Following a common practice in technology acceptance studies^31,32^, we used a case vignette to represent a typical situation in which an EHR app may be used. In particular, the case vignette depicted a situation where the participant has recently started using an EHR app and is now faced with the decision to upload a medical finding to their EHR (see Appendix). Additionally, the disease/injury was described in lay terms with one to three sentences (see Appendix). In selecting the diseases, both their stigmatization potential and their time course were systematically varied. The stigmatization potential covered different risks for professional and social life, such as depression as a disease^33,34^ or tests for STDs with high stigmatization potential^35,36^ and fractures or diabetes as diseases with low stigmatization potential. To reflect different time courses, diseases were divided according to an acute (e.g., wrist fracture) and a chronic time course (e.g., type-1 diabetes). Furthermore, diseases were selected to occur regardless of age so that they would be perceived as realistic diseases by an age-diverse sample. Table 2 shows the diseases used in the stimuli, categorized in terms of stigma potential and time course.

**Table 2.** Diseases used in the stimuli, categorized by stigmatization potential (SP) and time course (TC).

| SP \ TC | Acute | Chronic |
| --- | --- | --- |
| Low | Fractured wrist | Type-1 diabetes |
| High | STD (Gonorrhea) | Depression |

As in a previous study^16^, an interactive prototype (a so-called click dummy) was used which was modeled after the mobile EHR application of a German health insurance company (the BARMER) - the eCare app - using a software for interface design (FIGMA). This prototype allows for a realistic interaction with an EHR. Specifically, the prototype gave participants the ability to upload findings, grant or revoke permissions to view findings, and create medication plans. Only the “Upload findings” function was used in this study. The interaction with the EHR prototype preceded the survey study.

We used LimeSurvey (version 3.28.3+220315) to create and conduct a 5-page online survey. The EHR prototype was embedded into the survey using iFrame. LimeSurvey software was used to ensure that all questions had to be answered to complete the study and receive the compensation. As in the previous study, we tested the effect of the independent variables by querying the perceived risk and perceived benefit of uploading findings to the EHR.^16^ Based on previous results, we assumed a high perceived risk when the stigmatization potential was high and a high perceived benefit when the time course was chronic. Perceived risk, perceived benefit, and all questionnaire items^14^ (see Appendix) were measured using a 7-point Likert scale ranging from 1 (“Strongly disagree”) to 7 (“Strongly agree”).

### Procedure

The study procedure is shown in Figure 2. The survey consisted of 4 parts. After giving their informed consent according to the WMA Declaration of Helsinki, (1) participants had one minute to interact with the EHR prototype. (2a) Participants then read a randomly selected case vignette addressing the use of the EHR in the context of uploading a medical finding (low or high stigmatization potential and acute or chronic time course, depending on the experimental group). (2b) Subsequently, participants were asked to assess the risks and benefits of uploading the medical finding mentioned in the case vignette. (3) Afterwards the questionnaire was answered by the participants. (4) The survey was completed with the collection of demographic characteristics (age, gender, and education level) as control variables, as well as the opportunity for participants to declare their responses invalid due to lack of care in processing.

**Figure 2.**
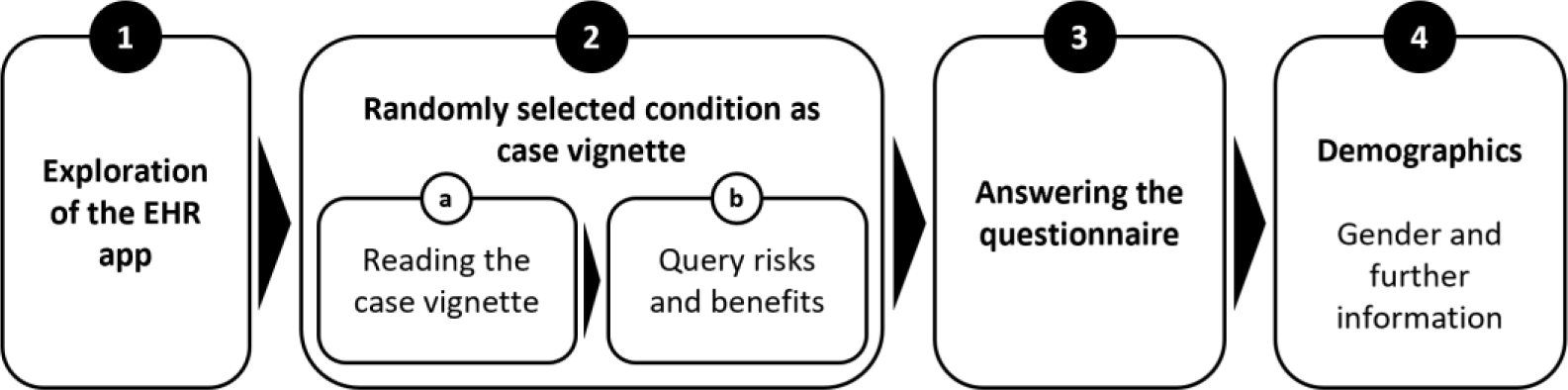
Overview of the study design.

**Figure 3.**
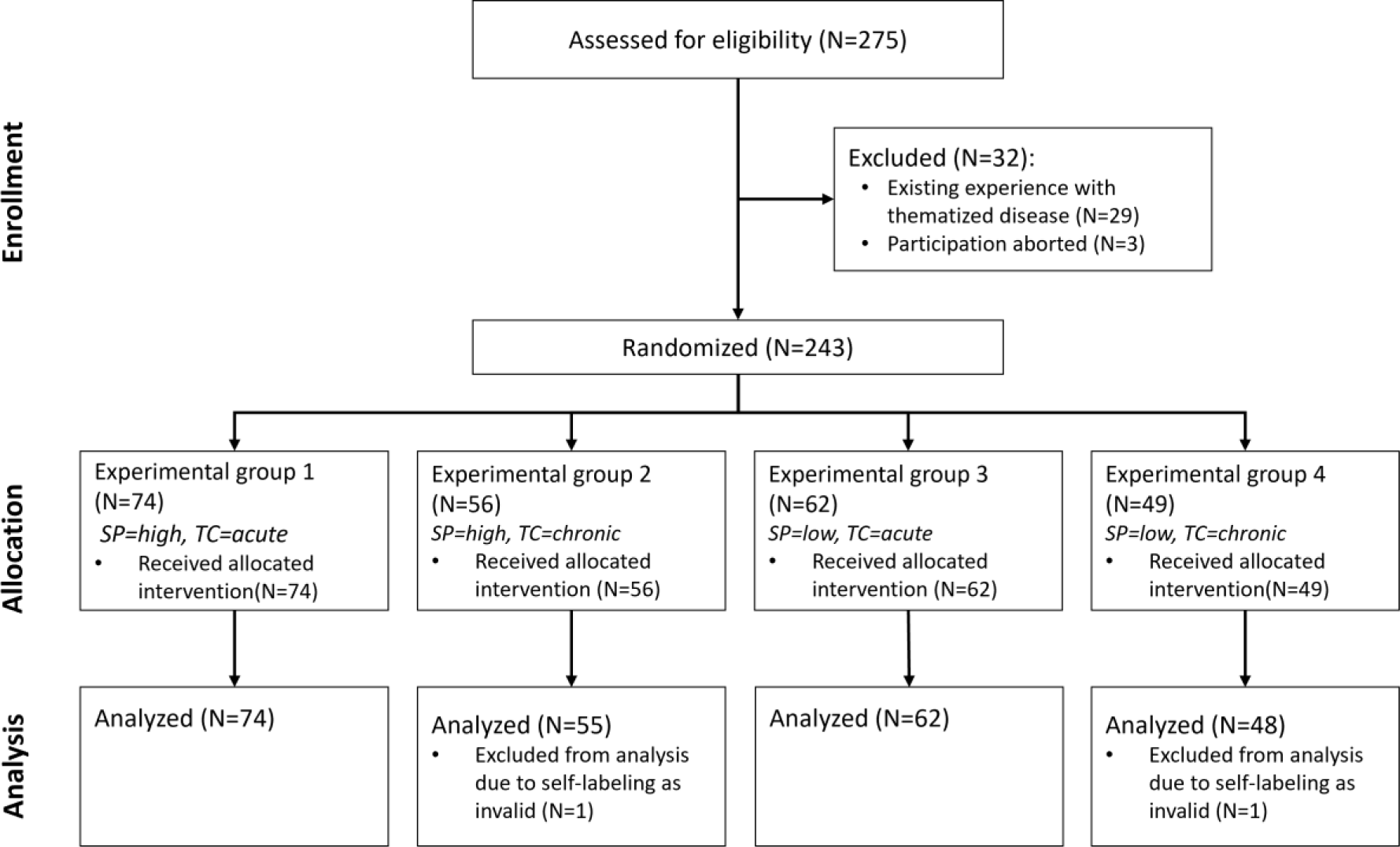
CONSORT Flow Chart. Note: SP = stigma potential, TC = time course.

### Analyses

To validate the model outlined in figure 1 and perform multigroup analyses (MGA) of the independent variables and controls, SEM-PLS was used. The SEM-PLS was carried out with SmartPLS 4 (version 4.0.9.4). All items of the questionnaire, except PB07 und SN03 due to poor standardized factor loadings, were included in the final analysis and restricted to load on the respective constructs describe above and in Figure 1. The *P* values of the paths were evaluated with t-statistics obtained from bootstrapping with 5,000 resamples. We cleaned and analyzed the data of the perceived risks and perceived benefits using RStudio (version 2023.06.0+421). These analyses were performed using t-tests.

## Results

### Survey characteristics

A total of 275 observations were collected. A total of 34 records were excluded, of which 29 were excluded because of participants’ medical history, three because of incomplete questionnaires, and two because they were marked as invalid by participants. Figure 2 shows the participation and distribution process of the participants according to the guidelines of the CONSORT statement.^37^

A sample of 241 observations (146 male, 92 female, 1 diverse, 2 no information) was used for further analysis. Table 3 summarizes the demographic characteristics of the sample.

**Table 3.** Demographic data of the sample (N=241).

| Demographic characteristic | Respondents |
| --- | --- |
| Age (years), mean (SD) | 31.31 (9.76) |
| **Gender, n (%)** |  |
| female | 92 (38.2) |
| male | 146 (60.6) |
| diverse | 1 (0.4) |
| no answer | 2 (0.8) |
| **Education, n (%)** |  |
| Highschool / vocational education | 120 (49.8) |
| Bachelor | 61 (25.3) |
| Master | 50 (20.7) |
| PhD | 9 (3.7) |
| other | 1 (0.4) |

### Risk and benefit perception

Similar to the preliminary study^16^, risk and benefit perception of uploading served as a manipulation check to test the validity of our manipulation of the stigmatization potential (risk) and the time course (benefit) through the medical findings (see Figure 4). As expected, uploading medical findings of stigmatized diseases was perceived as significantly riskier than those of non-stigmatized diseases (low: mean 3.98, SD 1.7; high: mean 5.12, SD 1.71; t_239_=5.447; *P*<.001). In addition, uploading findings of chronic diseases was perceived as significantly more beneficial than acute diseases (acute: mean 5.38, SD 1.37; chronic: mean 5.90, SD 1.37; t_239_=2.98, *P*=.003). Consequently, we assume that our manipulation was successful.

**Figure 4.**
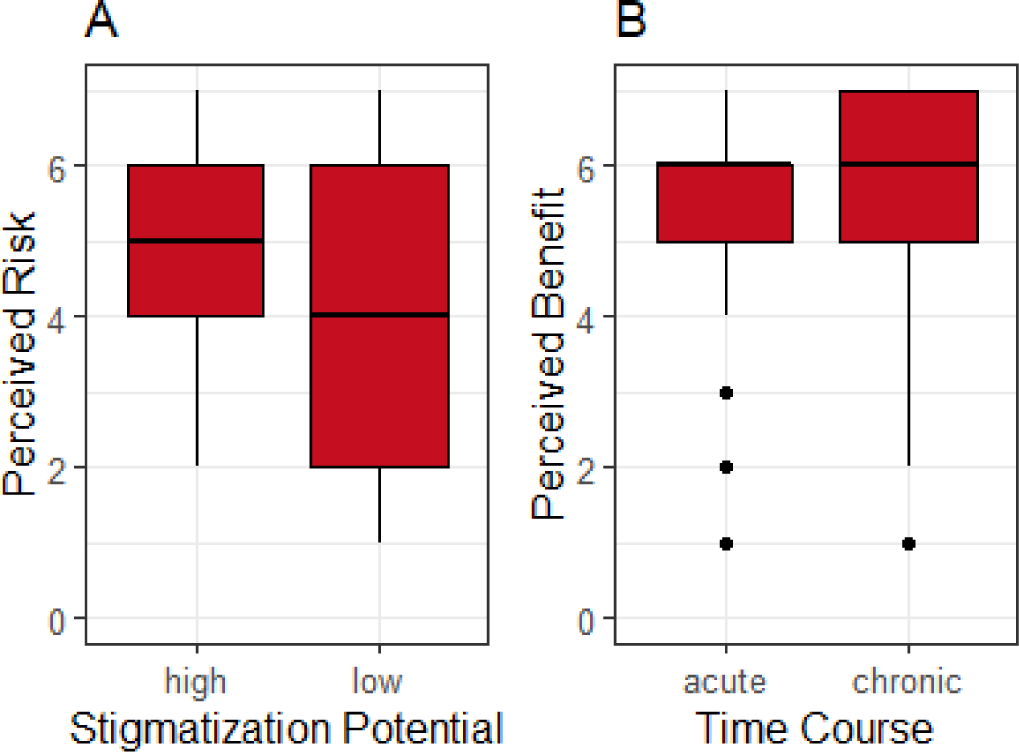
A) Perceived risk in terms of stigmatization potential (SP), and B) perceived benefit in terms of time course (TC).

### Assessment of the structural model

The internal consistency of the scales as well as convergent validity and discriminant validity of the measured constructs are shown in Table 4 and Table 5. Internal consistency was evaluated with Cronbach’s Alpha with the criterion of *α*≥.7.^38,39^ All constructs surpass the recommended value, so that internal consistency can be assumed. The convergent validity was assessed following Hair, Black, Babin and Anderson^39^ using three criteria: (1) The significance of the factor loadings, which exceed the criterion value of 0.5; (2) The Average Variance Extracted (AVE) should be greater than 0.5; (3) The Composite Reliability (CR) should surpass the minimum threshold of 0.6. After eliminating PB07 und SN03 due poor standardized factor loadings (<0.7)^39^, all subscales met these three criteria.

**Table 4.**
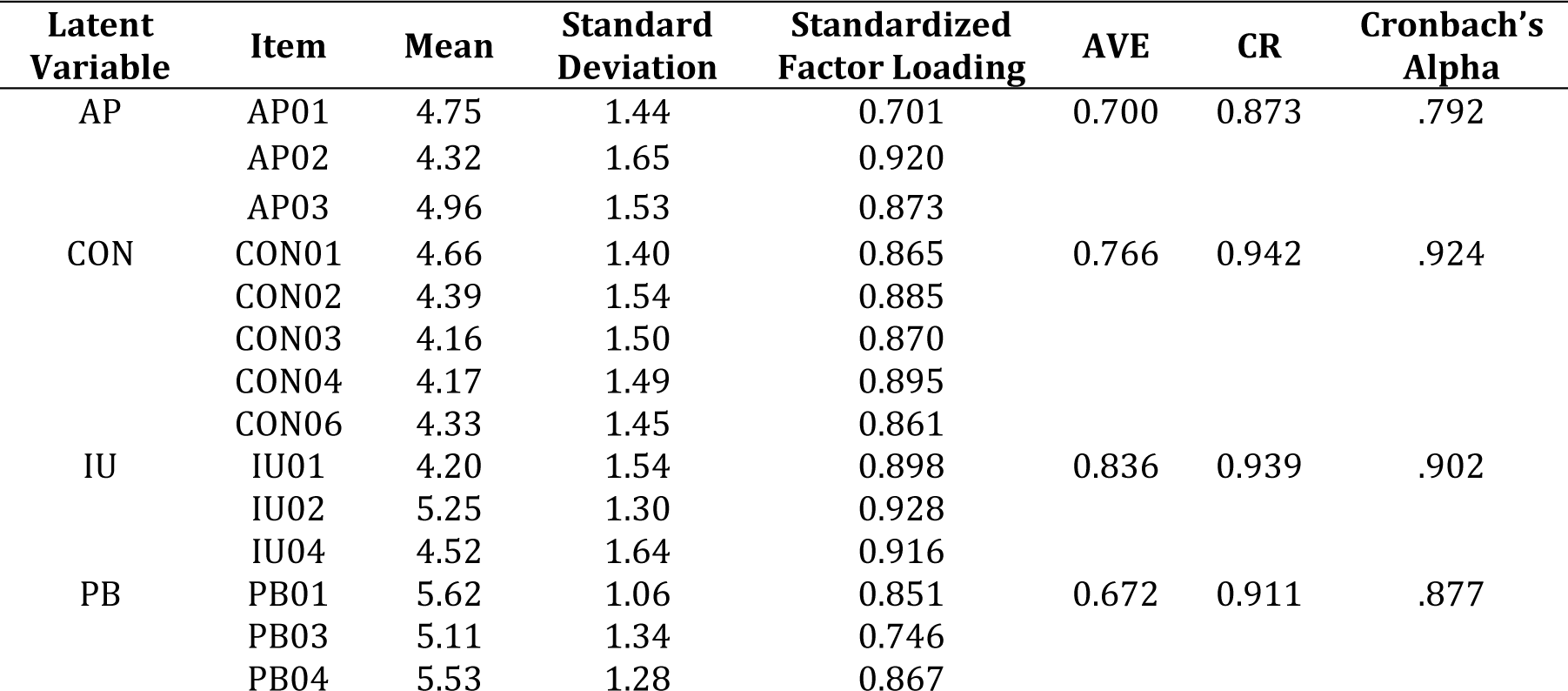
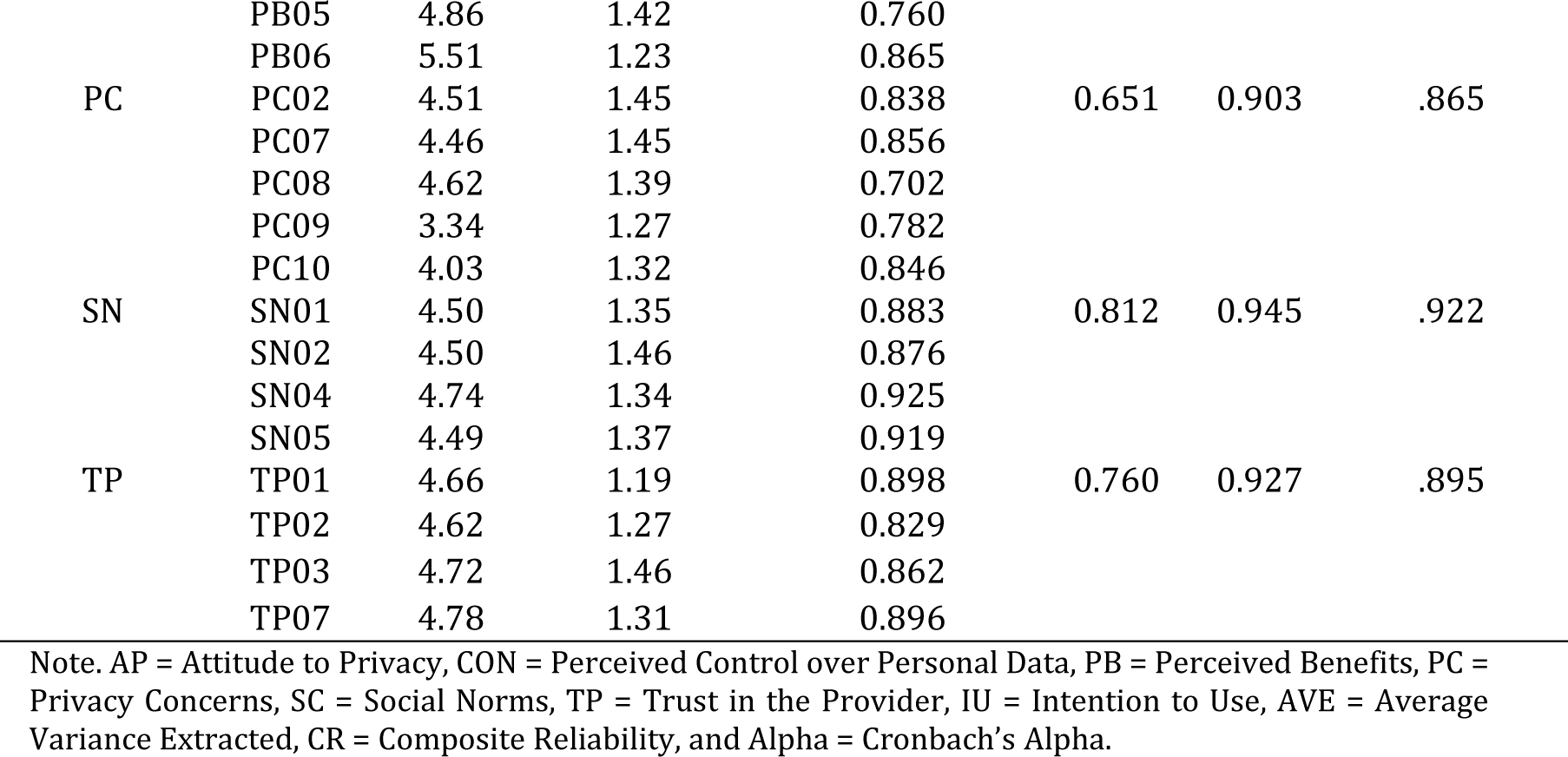
Quality criteria of the constructs.

| Latent Variable | Item | Mean | Standard Deviation | Standardized Factor Loading | AVE | CR | Cronbach's Alpha |
| --- | --- | --- | --- | --- | --- | --- | --- |
| AP | AP01 | 4.75 | 1.44 | 0.701 | 0.700 | 0.873 | .792 |
|  | AP02 | 4.32 | 1.65 | 0.920 |  |  |  |
|  | AP03 | 4.96 | 1.53 | 0.873 |  |  |  |
| CON | CON01 | 4.66 | 1.40 | 0.865 | 0.766 | 0.942 | .924 |
|  | CON02 | 4.39 | 1.54 | 0.885 |  |  |  |
|  | CON03 | 4.16 | 1.50 | 0.870 |  |  |  |
|  | CON04 | 4.17 | 1.49 | 0.895 |  |  |  |
|  | CON06 | 4.33 | 1.45 | 0.861 |  |  |  |
| IU | IU01 | 4.20 | 1.54 | 0.898 | 0.836 | 0.939 | .902 |
|  | IU02 | 5.25 | 1.30 | 0.928 |  |  |  |
|  | IU04 | 4.52 | 1.64 | 0.916 |  |  |  |
| PB | PB01 | 5.62 | 1.06 | 0.851 | 0.672 | 0.911 | .877 |
|  | PB03 | 5.11 | 1.34 | 0.746 |  |  |  |
|  | PB04 | 5.53 | 1.28 | 0.867 |  |  |  |
|  | PB05 | 4.86 | 1.42 | 0.760 |  |  |  |
|  | PB06 | 5.51 | 1.23 | 0.865 |  |  |  |
| PC | PC02 | 4.51 | 1.45 | 0.838 | 0.651 | 0.903 | .865 |
|  | PC07 | 4.46 | 1.45 | 0.856 |  |  |  |
|  | PC08 | 4.62 | 1.39 | 0.702 |  |  |  |
|  | PC09 | 3.34 | 1.27 | 0.782 |  |  |  |
|  | PC10 | 4.03 | 1.32 | 0.846 |  |  |  |
| SN | SN01 | 4.50 | 1.35 | 0.883 | 0.812 | 0.945 | .922 |
|  | SN02 | 4.50 | 1.46 | 0.876 |  |  |  |
|  | SN04 | 4.74 | 1.34 | 0.925 |  |  |  |
|  | SN05 | 4.49 | 1.37 | 0.919 |  |  |  |
| TP | TP01 | 4.66 | 1.19 | 0.898 | 0.760 | 0.927 | .895 |
|  | TP02 | 4.62 | 1.27 | 0.829 |  |  |  |
|  | TP03 | 4.72 | 1.46 | 0.862 |  |  |  |
|  | TP07 | 4.78 | 1.31 | 0.896 |  |  |  |

Note. AP = Attitude to Privacy, CON = Perceived Control over Personal Data, PB = Perceived Benefits, PC = Privacy Concerns, SC = Social Norms, TP = Trust in the Provider, IU = Intention to Use, AVE = Average Variance Extracted, CR = Composite Reliability, and Alpha = Cronbach's Alpha.

**Table 5.** Measurement model – Discriminant validity.

|  | AP | CON | IU | PB | PC | SN | TP |
| --- | --- | --- | --- | --- | --- | --- | --- |
| AP | **0.837** | 0.426 | 0.488 | 0.368 | 0.829 | 0.481 | 0.607 |
| CON | -0.411 | **0.875** | 0.689 | 0.649 | 0.727 | 0.586 | 0.841 |
| IU | -0.449 | 0.632 | **0.914** | 0.868 | 0.701 | 0.684 | 0.810 |
| PB | -0.338 | 0.593 | 0.782 | **0.820** | 0.534 | 0.590 | 0.721 |
| PC | 0.730 | -0.652 | -0.627 | -0.487 | **0.807** | 0.583 | 0.867 |
| SN | -0.428 | 0.543 | 0.625 | 0.536 | -0.527 | **0.901** | 0.719 |
| TP | -0.555 | 0.770 | 0.738 | 0.652 | -0.769 | 0.657 | **0.872** |

Notes: Fornell-Larcker: the diagonal elements (bold) are the square root of the variance shared between the constructs and their measures (average variance extracted); off-diagonal: the correlation between constructs; HTMT ratios are above the diagonal; AP = Attitude to Privacy, CON = Perceived Control over Personal Data, PB = Perceived Benefits, PC = Privacy Concerns, SC = Social Norms, TP = Trust in the Provider, and IU = Intention to Use.

Discriminant validity was evaluated by the Fornell–Larcker Criterion^39,40^. For each latent variable the square root of AVE (diagonal elements) must be larger than the correlation between this latent variable and any other latent variable (off-diagonal elements). Additionally, the heterotrait-monotrait (HTMT) ratio of correlations was used to certify discriminant validity^39,41^. All constructs were below the threshold of 0.90 for conceptual similar constructs. As shown in Table 5, both criteria were fulfilled for all latent variables. The measurement model thus presents appropriate psychometric properties.

### Results of the Structural Model

After evaluating the criteria of the measurement model, we now describe the structural model concerning the use of the EHR in more detail. The structural model was estimated by using bootstrapping (5,000 samples) to generate t-statistics and to evaluate the statistical significance of the relationships. Figure 5 represents the path coefficients and the corresponding *P* values.

**Figure 5.**
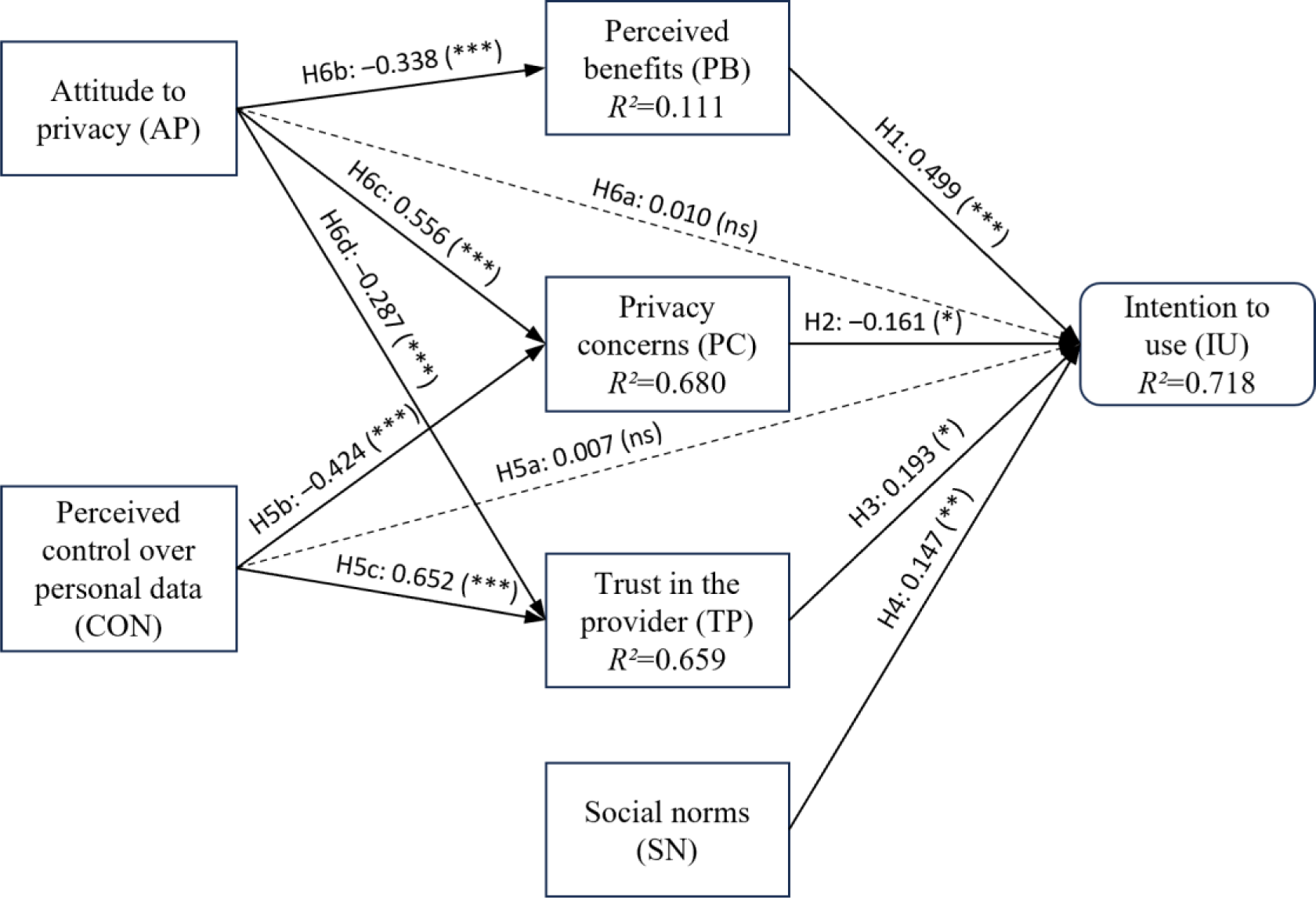
Factor relationships in the structural model. Solid lines represent statistically significant links and dashed lines represent statistically nonsignificant links. *P<.05. **P<.01. ***P<.001. ns: not significant

The *R²* values for the intention to use, privacy concerns and trust in the provider, but not for perceived benefits, far exceed the cutoff value of 0.4^42^ and suggests a good model fit. Our model explains *R²*=71.8% of the variance in our main dependent variable, the intention to use the EHR. Consistent with our expectations, perceived benefits have a significant effect on the intention to use (*P*<.001), as well as privacy concerns (*P*=.018), trust in the provider (*P*=.010) and social norms (*P*=.001), supporting H1, H2, H3 and H4. Perceived control over personal data has significant effects on privacy concerns (*P*<.001) and trust in the provider (*P*<.001), thus supporting H5b and H5c. There is no significant effect on intention to use (*P*=.897), rejecting H5a. The attitude to privacy has significant effects on perceived benefits (*P*<.001), privacy concerns (*P*<.001) and trust in the provider (*P*<.001). Thus, H6b, H6c and H6d are supported. The attitude to privacy, however, has no effect on the intention to use (*P*=.841). Consequently, H6a is rejected. Table 6 summarizes the detailed analysis of the path coefficients.

**Table 6.** Overview of the path coefficients.

| Hypotheses | Path | Path coefficient | t value | P value | Supported? |
| --- | --- | --- | --- | --- | --- |
| H1 | PB → IU | +0.499 | 9.526 | <.001 | Yes |
| H2 | PC → IU | -0.161 | 2.359 | .018 | Yes |
| H3 | TP → IU | +0.193 | 2.592 | .010 | Yes |
| H4 | SN → IU | +0.147 | 3.362 | <.001 | Yes |
| H5a | CON → IU | +0.007 | 0.129 | .897 | No |
| H5b | CON → PC | -0.424 | 10.353 | <.001 | Yes |
| H5c | CON → TP | +0.652 | 18.008 | <.001 | Yes |
| H6a | AP → IU | +0.010 | 0.200 | .841 | No |
| H6b | AP → PB | -0.338 | 5.797 | <.001 | Yes |
| H6c | AP → PC | +0.556 | 15.265 | <.001 | Yes |
| H6d | AP → TP | -0.287 | 7.315 | <.001 | Yes |

### Results of the Multigroup Analysis (MGA)

We conducted multigroup analyses (MGA) to examine whether the relationships in the model varied depending on the control variables (i.e., personal characteristics) and the IVs. The MGA allows for direct nonparametric tests of the path estimates in the structural model for each bootstrap sample, following the MGA procedure in PLS with a bootstrapping of 5,000 samples.^43^

### Controls

Aside from gender (female vs. male), we recoded the demographic variables age (younger vs. older) and education (non-academics vs. academics) as dichotomous variables, because MGA are performed as two-group comparisons. The influence of gender (female: 92 cases; male: 146 cases) showed no significant difference in the path coefficients. For age, we divided the participants into two age groups by performing a median split, younger adults (participants 18-29 years of age: 128 cases) and older adults (participants who are 30 years or older: 113 cases). There were significant differences in the path coefficients between the younger and older participants regarding the influence of privacy concerns (*P*=.034) and trust in the provider (*P*=.016) on intention to use. While privacy concerns do not influence intention to use for younger participants (path coefficient −0.003, *P*=.980), their influence increases with increasing age (path coefficient −0.280, *P*=.001). For the younger participants, trust in the provider influenced the intention to use (path coefficient 0.315, *P*<.001), although not for the older participants (path coefficient - 0.012, *P*=.907). For education (non-academics: 120 cases; academics: 121 cases) there were significant differences for the path coefficients between trust in the provider (*P*=.008) and intention to use. Trust in the provider influences the intention to use the service for non-academics (path coefficient 0.384, *P*<.001) but not for academics (path coefficient 0.011, *P*=.910).

### Time course and stigmatization potential

The MGA for the stigmatization potential of diseases (low: 111 cases; high: 130 cases) showed no significant differences in the path coefficients. However, there were significant differences with respect to the time course of diseases (acute: 136 cases; chronic: 105 cases) between the path estimates between the influence of perceived benefits (*P*=.008), privacy concerns (*P*=.025), social approval (*P*=.011), trust in the provider (*P*=.012), attitude to privacy (*P*=.050) and perceived control over personal data (*P*=.012) on intention to use (see Figure 6).

**Figure 6.**
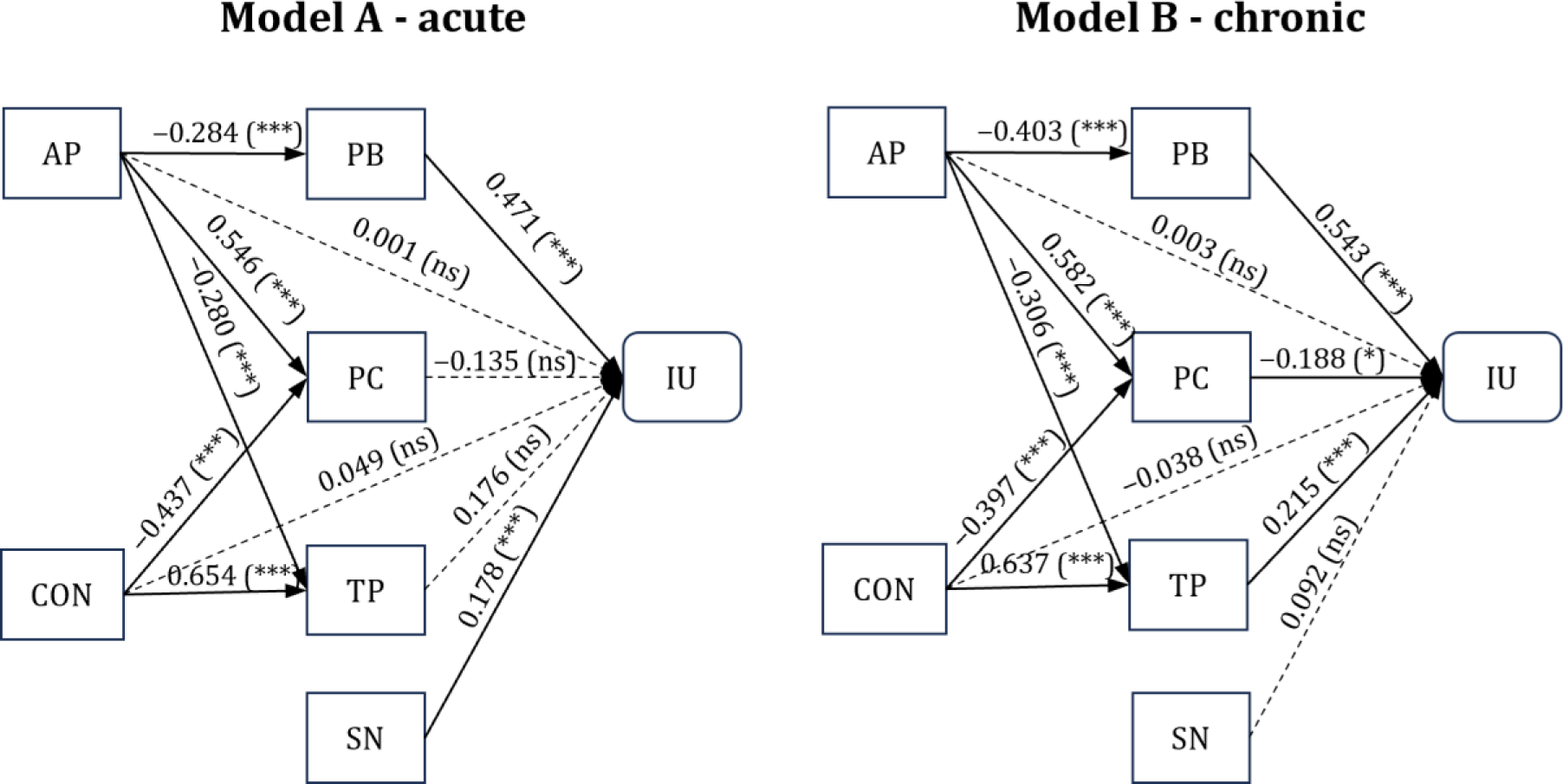
Factor relationships in the structural sub-models concerning acute and chronic diseases. Solid lines represent statistically significant links and dashed lines represent statistically nonsignificant links. Notes: *P<.05. **P<.01. ***P<.001. ns: not significant; AP = Attitude to Privacy, CON = Perceived Control over Personal Data, PB = Perceived Benefits, PC = Privacy Concerns, SN = Social Norms, TP = Trust in the Provider, and IU = Intention to Use.

When uploading medical findings to the EHR, the perceived benefits had a greater influence on intention––albeit in the same direction––when the mentioned disease was chronic (path coefficient 0.543, *P*<.001), than in the case of an acute disease (path coefficient 0.471, *P*<.001). When interacting with medical reports of chronic diseases, privacy concerns influence intention to use (path coefficient −0.188, *P*=.049) in contrast to acute diseases (path coefficient −0.135, *P*=.138). Conversely, social approval had no influence on intention to use when uploading reports of chronic diseases (path coefficient 0.092, P=.244), but they did when uploading reports of acute diseases (path coefficient 0.178, P=.001). When uploading reports of chronic diseases, trust in the provider had an influence on intention to use (path coefficient 0.215, *P*=.039), but not when uploading findings of acute diseases (path coefficient 0.176, *P*=.098). Although there was a significant difference for uploading acute and chronic findings to the EHR in terms of the influence of both perceived control over personal data and attitudes towards privacy on intention to use, the influence of both paths was not significant in both sub-models and therefore of no further importance. Table 7 presents the path coefficients and the corresponding P-values of the models of the subsamples with respect to the time course as well as the difference between the models.

**Table 7.** Results of the MGA concerning time course.

| Path | Path coefficients (acute) | P value (acute) | Path coefficients (chronic) | P value (chronic) | Path coefficients (difference) | P value (difference) |
| --- | --- | --- | --- | --- | --- | --- |
| PB → IU | 0.471 *** | <.001 | 0.543 *** | <.001 | 0.072 *** | .008 |
| PC → IU | -0.135 ns | .138 | -0.188 * | .049 | -0.053 * | .025 |
| TP → IU | 0.176 ns | .098 | 0.215 * | .039 | 0.039 * | .012 |
| SN → IU | 0.178 *** | .001 | 0.092 ns | .244 | -0.086 * | .011 |
| CON → IU | 0.049 ns | .544 | -0.038 ns | .623 | -0.087 * | .012 |
| CON → PC | -0.437 *** | <.001 | -0.397 *** | <.001 | 0.04 ns | .618 |
| CON → TP | 0.654 *** | <.001 | 0.637 *** | <.001 | -0.017 ns | .565 |
| AP → IU | 0.001 ns | .989 | 0.003 ns | .966 | 0.002 ns | .050 |
| AP → PB | -0.284 *** | <.001 | -0.403 *** | <.001 | -0.119 ns | .365 |
| AP → PC | 0.546 *** | <.001 | 0.582 *** | <.001 | 0.036 ns | .561 |
| AP → TP | -0.280 *** | <.001 | -0.306 *** | <.001 | -0.026 ns | .795 |

## Discussion

### Principal Findings

This study investigated whether the privacy calculus model adapted for mHealth apps can describe the intention to use the EHR in general and in relation to personal characteristics (demographics) and disease characteristics including stigmatization potential and time course. The model provides a valid representation of the intention to use the EHR, as the proposed model explains the variance (R²) in users’ intention to use mHealth applications better than other reported models (where the values do not exceed 0.5)^15,44^, with an overall complexity similar to existing models. We examined the influence of demographic control variables on the relationships in the model, of which age and education had significant effects. Older users have more privacy concerns and less trust in the provider than younger users, which negatively influences their intention to use the EHR. Additionally, the influence of trust in the provider on the intention to use the EHR also decreased as the level of education increased. While the influence of both control variables is not uncommon in the mHealth and IoT domain^25,26^, they did not appear in other SEM studies regarding EHR use thus far.^10,15,45^

When looking at the full model, it is evident that the intention to use the EHR is significantly influenced by perceived benefits (path coefficient=0.499) as the overall strongest driver of intention to use. The more beneficial the use of the EHR is perceived by users, e.g., for health data management of patients with multiple chronic conditions,^46^ the more likely it is that an intention to use it will be formed. However, these perceived benefits were inhibited by the patient’s attitude to privacy (negative path coefficient=-0.338). This means that benefits of EHR use, such as more effective treatment and less effort in managing one’s health data, are devalued or ignored in the decision to use the EHR if the respective patients perceive privacy issues as important. The inhibiting influence of privacy concerns on the intention to use (negative path coefficient=-0.161) confirms similar findings of prior studies.^7^ Privacy concerns were even more pronounced when general attitudes to privacy were high (path coefficient = 0.556). Nevertheless, our model suggests that privacy concerns were significantly inhibited by the level of perceived control over personal data (negative path coefficient=-0.424). The more users perceive that they can make independent decisions about whether and how to store their data in the EHR (delete, share, etc.), the fewer privacy concerns they tend to have.^45^ The model also shows that the more trust is placed in the provider, the more likely it is that the EHR will be used (path coefficient=0.193). This trust is positively associated with control over personal data (path coefficient=0.652) unless patients have a generally high attitude to privacy (negative path coefficient = 0.287). Finally, the intention to use was also driven by social norms (i.e., social approval), that is, the opinions, experiences, and the EHR use of close relatives (path coefficient=0.147). This is in line with findings from social psychology, which show that people tend to adopt the opinions of their peers or relatives.^47^

The consideration of disease characteristics (time course and stigmatization potential) enable a more nuanced view of the factors influencing the intention to use the EHR. An important finding is that the intention to use the EHR is significantly influenced by the time course of diseases but not the stigmatization potential, which has been shown to influence actual uploading behavior.^16,17^

For acute diseases, the intention to use EHR is primarily influenced by the perceived benefits (path coefficient=0.471) and social approval (path coefficient=0.178). In the case of chronic diseases, the situation is more complex. Here, the intention to use EHR is affected by three key factors: perceived benefits (path coefficient=0.543), privacy concerns (negative path coefficient=-0.188), and trust in the provider (path coefficient= 0.215). In other words, the significant differences regarding the acute and chronic time course of diseases can be broken down into four main points regarding perceived benefits (a), privacy concerns (b) trust in the provider (c) and social norms (d):

a. When comparing the impact of perceived benefits on the intention to use the EHR, their positive influence is notably stronger in the case of chronic diseases than it is for acute diseases. This is not surprising as health concerns (i.e., the personal importance of health issues) have a positive impact on the sharing of health data via the EHR.^10^ It is in one’s own health interest to share data about chronic diseases via the EHR so that it can be considered in future diagnoses.^10,48^
b. The impact of privacy concerns varies significantly between acute and chronic diseases. Although privacy concerns tend to decrease intentions to use the EHR in the case of chronic diseases, they do not significantly influence the intention to use EHR with acute diseases.^7^ This finding is particularly striking given that previous studies suggest that patients had less privacy concerns when the use of an EHR meant an improvement in their health.^10,49^ Contrary to this, our observation indicates an increased influence of privacy concerns in chronic diseases, i.e. when EHR use would result in an actual improvement in users’ health.
c. Trust in the provider had a positive influence on the intention to use the EHR only in the case of chronic diseases. This is hardly surprising, as a perceived vulnerability (i.e., awareness of a possible risk) is a prerequisite for trust. If little risk is perceived, as with the acute diseases in our study, no trust is required to upload the findings to the EHR.^50,51^
d. For chronic diseases the influence of social norms on the intention to use the EHR vanished. Consequently, when it comes to chronic diseases patients intention to the EHR is neither influenced by the approval of their social group (i.e.,. injunctive norms or what we believe others think we should do) nor by the behavior of others (i.e., descriptive norms or typical behavior in a situation). Instead, they seem to rely on their own perception of benefits and risks.

### Implications

Our study has various theoretical and practical implications. As can be seen from the differences between the sub-models in terms of the time course of the disease as wells as age and education of users, intention to use the EHR is a context-specific construct. The focus on context increases the usefulness of these models as it helps to further specify which factors should be considered when conducting studies with the privacy calculus model and how to translate its predictions into practical interventions to increase EHR adoption rates. Specifically, social norms / approval may be effective to increase EHR uptake when it comes to acute diseases (e.g., through a reward system for encouraging family members or friends) but not for chronic diseases. For chronic diseases as well as for older and higher educated users, on the other side, privacy concerns should be reduced and trust in the provider should be built, for instance, through transparent and easy-to-understand information about the high safety standards of EHR (e.g., for encrypting data) and the data stored in them (e.g., via the control of access rights). Such combined measures can help to increase the number of users, regardless of disease characteristics and thus support implementation efforts of EHR.

### Limitations and Future Directions

We deliberately excluded participants who already had a medical history with the diseases addressed in the stimuli to avoid bias in their responses. Individuals suffering from a stigmatized disease are more cautious to disclose the information, especially if the disease is not immediately apparent.^28,52^ The question arises as to what extent the behavior of stigmatized and chronically ill people can be simulated under experimental conditions, provided that the participants do not have any stigmatized characteristics or chronic illnesses. To further strengthen the validity and generalizability of our results, the perspective of people who are already affected should be examined in a follow-up study and compared with the results of this study. Another limitation is that the chronic and acute disease patterns used in the stimuli are not readily comparable. We decided to use the diseases listed in Table 1 because they resulted in the expected effects in a preliminary study.^16^ For future studies, it would make sense to use other diseases that can be more readily compared in terms of their perceived stigma and time course (e.g., gonorrhea and HIV or a wrist fracture and arthritis) to further strengthen the generalizability of the present findings. In addition, the study was conducted exclusively with German citizens due to the focus on the German healthcare system. Follow-up studies should examine the applicability of the results to other countries, as there are differences between the individual healthcare systems as well as in the willingness to share personal data online.^53^ Finally, in this study, injunctive social norms were operationalized with respect to recommendations and approval of the use of the EHR by friends and families. To what extent patient’s beliefs of social approval of health professionals increase or decrease intention to use^54^ remains to be seen in future studies.

### Conclusions

This paper highlights that disease and personal characteristics influence the formation of the intention to use EHRs. Specifically, the study found relevant differences in the intention to use based on the time course of diseases to be uploaded in the EHR. In the case of acute illnesses, the intention to use the EHR was primarily driven by perceived benefits and social norms. For chronic diseases, the study observed a significant shift. The influence of social norms on the intention to use the EHR vanished, whereas factors like privacy concerns, trust in the provider, and perceived benefits became more prominent. This suggests that patients with chronic illnesses recognize a greater value in using the EHR, but they have heightened privacy concerns due to the more permanent and risky nature of the data associated with chronic conditions. Additionally, the study found that the experiences, recommendations, and behavior of close relatives influenced the intention to use the EHR only for acute diseases. Finally, personal characteristics also had an influence on the intention to use the EHR. With increasing levels of education, trust in the provider no longer played a role in the formation of the intention to use. Similarly, compared to their younger counterparts, older users’ intention to use the HER was less influenced by trust in their provider and more impacted by privacy concerns.

These findings imply that for effective EHR adoption, strategies must be tailored to the specific needs and concerns of different patients with different types of diseases. This more nuanced understanding can guide healthcare providers and policymakers in developing targeted approaches to increase user adoption, ensuring that the system meets the varying needs of patients with different health conditions. This is an important step for a more successful and widespread implementation of the EHR.

## Data Availability

All data produced in the present study are available upon reasonable request to the authors.

## Acknowledgements

We thank all those who participated in the study.

## Conflicts of interest

The author(s) declared no potential conflicts of interest with respect to the research, authorship, and/ or publication of this article.

## Ethical approval

This Ethics Committee of the Department of Psychology and Ergonomics (Institut für Psychologie und Arbeitswissenschaft [IPA]) at Technische Universität Berlin approved this study (tracking number: AWB_KAL_1_230311).

## Funding

The authors received no financial support for the research and/or authorship of this article. We acknowledge support from the German Research Foundation and the Open Access Publication Fund of TU Berlin. We also thank the evangelisches Studienwerk Villigst and the German Federal Ministry of Education and Research, who provided the doctoral scholarship (NvK) without which this research would not have been possible.

## Guarantor

NvK

## Contributorship

NvK researched literature and conceived the study under the supervision of MF. NvK was involved in protocol development, gaining ethical approval, patient recruitment and data analysis. NvK wrote the first draft of the manuscript. All authors reviewed and edited the manuscript and approved the final version of the manuscript.

## Appendix

### Case vignettes

#### Case vignette 1

1. you have recently started using the electronic health record (EHR), which you have just learned about, to manage your medical data and records and to share them with your physicians if you wish.
2. you have recently contracted a sexually transmitted disease (gonorrhea) but have completely recovered.

You are now faced with the decision of whether or not to include the findings of this disease in your EHR.

#### Case vignette 2

1. you have recently started using the electronic health record (EHR), which you have just learned about, to manage your medical data and records and to share them with your physicians if you wish.
2. you have been suffering from moderate to severe depression for several years.

You are now faced with the decision of whether or not to include the findings of this disease in your EHR.

#### Case vignette 3

1. you have recently started using the electronic health record (EHR), which you have just learned about, to manage your medical data and records and to share them with your physicians if you wish.
2. You recently broke your wrist, but it has healed completely.

You are now faced with the decision of whether or not to include the findings of this disease in your EHR.

#### Case vignette 4

1. you have recently started using the electronic health record (EHR), which you have just learned about, to manage your medical data and records and to share them with your physicians if you wish.
2. You have had diabetes for several years.

You are now faced with the decision of whether or not to include the findings of this disease in your EHR.

## Description of the diseases

### Gonorrhea

Gonorrhea (gonorrhea) is an infectious disease that is mainly transmitted during sex. The pathogens are bacteria known as gonococci. Gonorrhea is one of the most common sexually transmitted diseases, also known as STDs. Gonorrhea can usually be treated well with antibiotics. If it remains untreated, complications are possible - such as joint inflammation, abdominal pain or fertility problems.

### Depression

Depression is a mental illness that can manifest itself in numerous symptoms. A persistently depressed mood, inhibition of drive and thinking, loss of interest and a variety of physical symptoms, ranging from insomnia to appetite disorders and pain, are possible signs of depression. The majority of those affected have suicidal thoughts sooner or later, and 10 to 15% of all patients with recurring severe depressive phases die by suicide. Once the correct diagnosis has been made, the situation is anything but hopeless. In recent decades, a lot has been done in terms of treatment and more than 80% of sufferers can be helped permanently and successfully.

### Wrist fracture

A wrist fracture is a fracture of the radius (one of the two forearm bones) close to the wrist. The medical term is “distal radius fracture”. Wrist fractures are the most common form of bone fracture in adults. A wrist fracture often heals without any problems, especially in the case of stable fractures. In very rare cases, complications and late effects develop, such as limited mobility or loss of strength in the wrist and fingers.

### Diabetes

Type 1 diabetes is an autoimmune disease in which the patient’s own immune system attacks the body’s own insulin production in the pancreas and destroys the insulin-producing cells: this results in an absolute insulin deficiency, which leads to a sharp rise in blood sugar and a simultaneous undersupply of the body’s cells. Lifelong therapy with insulin injections is therefore necessary, as well as adapting the diet to the insulin dosage in order to prevent blood sugar fluctuations. Accompanying symptoms are often other autoimmune diseases.

### Questionnaire

**Manipulation check (perceived risk):** Rate the following statement: If the information in this finding fell into the wrong hands, it would do me a lot of damage.

**Manipulation check (perceived benefit):** Rate the following statement: I think it is very beneficial that physicians who treat me have access to this finding through the electronic health record (EHR).

### Attitude to privacy

**AP01**: Compared to other issues that concern me, privacy is very important to me.

**AP02**: I feel very concerned about my privacy when using data-collecting technology.

**AP04**: I worry about what data is being stored about me.

### Perceived control over personal data

**CON01**: I believe I can control my personal data provided to the EHR.

**CON02**: I believe I have control over who can get access to my personal data collected by the EHR.

**CON03**: I think I have control over what personal information is released by the provider of the EHR.

**CON04**: I believe I have control over how personal data is used by the provider of the EHR.

**CON06**: Privacy settings allow me to have full control over the data I provide.

### Intention to use

**IU01**: I should use the EHR as soon as possible.

**IU02**: I would use the EHR.

**IU04**: I would not hesitate to use the EHR.

### Perceived benefits

**PB01**: Providing my personal data to the EHR will entail benefits.

**PB03**: I believe that as a result of the upload of my health date to the EHR, I will benefit from a better, more customized service.

**PB04**: I feel that uploading my health data to the EHR is useful.

**PB05**: Uploading my health data to the EHR can make my lifestyle more productive.

**PB06**: Uploading my health data to the EHR can simplify my lifestyle and accelerate processes.

**PB07**: The usage of the EHR can lead me to learn new things or think about things in new ways.

### Privacy concerns

**PC02**: I am concerned about submitting data to the EHR, because it could be used in a way I did not foresee.

**PC07**: There would be high potential for privacy loss associated with giving personal data to the EHR.

**PC08**: Personal data could be inappropriately used by the EHR.

**PC09**: Providing the EHR with my health data would involve many unexpected problems.

**PC10**: It would be risky to give health data to the EHR.

### Social norms

**SN01:** If my family recommends that I use the EHR, I do.

**SN02**: If my social environment used the EHR, I wouldn’t have any problems using it either.

**SN03:** I feel that I should use the EHR because everybody else seems to be using it.

**SN04**: If people I value, recommend me to use the EHR, I would.

**SN05**: If my friends were using the EHR, so would I.

### Trust in the provider

**TP01**: The provider of the EHR would be trustworthy in handling the data.

**TP02**: The provider of the EHR would tell the truth and fulfill promises related to the data provided by me.

**TP03**: I trust that the provider of the EHR would keep my best interests in mind when dealing with the data.

**TP07**: I would trust the EHR.

### Demographics

**Age**: Please enter your age.

**Gender (m/f/d):** Please indicate your gender.

**Education**: Please enter your highest qualification.

**Experience with mHealth apps**: How often do you use health or fitness apps?

## Abbreviations

AP: attitude to privacy
AVE: average variance extracted
CON: perceived control over personal data
CONSORT: consolidated standards of reporting trails
CR: composite reliability
EHR: electronic health record
HTMT: heterotrait-monotrait ratio of correlations
IoT: internet of things
IU: intention to use
IV: independent variable
M: mean
MGA: multigroup analysis
mHealth: mobile health
OR: odds ratio
PB: perceived benefits
PC: privacy concerns
R²: coefficient of determination
SD: standard deviation
SEM: structural equation modeling
SEM-PLS: structural equation modeling with partial least squares
SN: social norm
SNS: social network site
SP: stigmatization potential
STD: sexually transmitted disease
TC: time course
TP: trust in the provider

## Notes

### Competing Interest Statement

The authors have declared no competing interest.

### Funding Statement

The authors received no financial support for the research and/or authorship of this article. We also thank the evangelisches Studienwerk Villigst and the German Federal Ministry of Education and Research, who provided the doctoral scholarship (NvK) without which this research would not have been possible.

### Author Declarations

This Ethics Committee of the Department of Psychology and Ergonomics (Institut fuer Psychologie und Arbeitswissenschaft [IPA]) at Technische Universitaet Berlin approved this study (tracking number: AWB_KAL_1_230311).

## References

1. Galetsi P, Katsaliaki K, Kumar S. Values, challenges and future directions of big data analytics in healthcare: A systematic review. Social Science & Medicine 2019; 241: 112533.

2. Haleem A, Javaid M, Pratap Singh R, et al. Medical 4.0 technologies for healthcare: Features, capabilities, and applications. Internet of Things and Cyber-Physical Systems 2022; 2: 12–30.

3. Bertram N, Püschner F, Gonçalves ASO, et al. Einführung einer elektronischen Patientenakte in Deutschland vor dem Hintergrund der internationalen Erfahrungen. In: Klauber J, Geraedts M, Friedrich J, et al. (eds) Krankenhaus-Report 2019. Berlin, Heidelberg: Springer Berlin Heidelberg, pp. 3–16.

4. Bundesministerium für Gesundheit (BMG). Die elektronische Patientenakte (ePA), https://www.bundesgesundheitsministerium.de/elektronische-patientenakte.html (2021, accessed 14 April 2023).

5. Baron von Osthoff M, Watzlaw-Schmidt U, Lehmann T, et al. Patientengruppenspezifische Datenhoheitsbedürfnisse und Akzeptanz der elektronischen Patientenakte. Bundesgesundheitsbl. Epub ahead of print 23 September 2022. DOI: 10.1007/s00103-022-03589-w.

6. Hubmann M, Pätzmann-Sietas B, Morbach H. Telemedizin und digitale Akte – Wo stehen wir?: Chancen und Herausforderungen bei der Umsetzung in Klinik-und Praxisalltag. Monatsschr Kinderheilkd 2021; 169: 711–716.

7. Kus K, Kajüter P, Arlinghaus T, et al. Die elektronische Patientenakte als zentraler Bestandteil der digitalen Transformation im deutschen Gesundheitswesen – Eine Analyse von Akzeptanzfaktoren aus Patientensicht. HMD 2022; 59: 1577–1593.

8. Nøhr C, Parv L, Kink P, et al. Nationwide citizen access to their health data: analysing and comparing experiences in Denmark, Estonia and Australia. BMC Health Serv Res 2017; 17: 534.

9. Hertzum M, Ellingsen G, Cajander Å. Implementing Large-Scale Electronic Health Records: Experiences from implementations of Epic in Denmark and Finland. International Journal of Medical Informatics 2022; 167: 104868.

10. Cherif E, Bezaz N, Mzoughi M. Do personal health concerns and trust in healthcare providers mitigate privacy concerns? Effects on patients’ intention to share personal health data on electronic health records. Social Science & Medicine 2021; 283: 114146.

11. Taylor J, Corderoy A. My Health Record: almost $2bn spent but half the 23m records created are empty. The Guardian, 22 January 2020, https://www.theguardian.com/australia-news/2020/jan/23/my-health-record-almost-2bn-spent-but-half-the-23m-records-created-are-empty (22 January 2020, accessed 27 November 2023).

12. Bertelsmann Stiftung. Studie - Elektronische Patientenakte: Geplante Widerspruchslösung trifft auf breite Zustimmung, https://www.bertelsmann-stiftung.de/de/themen/aktuelle-meldungen/2023/februar/elektronische-patientenakte-geplante-widerspruchsloesung-trifft-auf-breite-zustimmung?tx_rsmbstpress_pi1%5Bpage%5D=1&cHash=d31de4f981340aa76e0265c48fa79163 (2023, accessed 17 May 2023).

13. Bundesministerium für Gesundheit (BMG). Patientendaten-Schutz-Gesetz, https://www.bundesgesundheitsministerium.de/patientendaten-schutz-gesetz.html (2020, accessed 14 April 2023).

14. Von Kalckreuth N, Feufel MA. Extending the Privacy Calculus to the mHealth Domain: Survey Study on the Intention to Use mHealth Apps in Germany. JMIR Hum Factors 2023; 10: e45503.

15. Dinev T, Albano V, Xu H, et al. Individuals’ Attitudes Towards Electronic Health Records: A Privacy Calculus Perspective. In: Gupta A, Patel VL, Greenes RA (eds) Advances in Healthcare Informatics and Analytics. Cham: Springer International Publishing, pp. 19–50.

16. von Kalckreuth N, Prümper AM, Feufel MA. The Influence of Health Data on the Use of the Electronic Health Record (EHR) – a Mixed Methods Approach. In: AMCIS 2023 Proceedings. 2023.

17. Von Kalckreuth N, Feufel MA. Acceptance ≠ Usage: what influences patients’ decisions to upload medical reports to the electronic health record (EHR)? A randomized controlled trial in Germany (Preprint). Preprint, JMIR Human Factors. Epub ahead of print 10 September 2023. DOI: 10.2196/preprints.52625.

18. Naeem I, Quan H, Singh S, et al. Factors Associated With Willingness to Share Health Information: Rapid Review. JMIR Hum Factors 2022; 9: e20702.

19. Kokolakis S. Privacy attitudes and privacy behaviour: A review of current research on the privacy paradox phenomenon. Computers & Security 2017; 64: 122–134.

20. Kim B, Kim D. Understanding the Key Antecedents of Users’ Disclosing Behaviors on Social Networking Sites: The Privacy Paradox. Sustainability 2020; 12: 5163.

21. Xu H, Dinev T, Smith HJ, et al. Examining the Formation of Individual’s Privacy Concerns: Toward an Integrative View. ICIS 2008 Proceedings 2008; 6: 1–17.

22. Xu F, Michael K, Chen X. Factors affecting privacy disclosure on social network sites: an integrated model. Electron Commer Res 2013; 13: 151–168.

23. von Kalckreuth N, Feufel MA. Disclosure of Health Data – Conceptualizing the Intention to use Wearables as an Extended Privacy Calculus. In: AMCIS 2021 Proceedings. 2021, pp. 1–6.

24. Schomakers E-M, Lidynia C, Ziefle M. Exploring the Acceptance of mHealth Applications - Do Acceptance Patterns Vary Depending on Context? In: Ahram TZ (ed) Advances in Human Factors in Wearable Technologies and Game Design. Cham: Springer International Publishing, pp. 53–64.

25. Uncovska M, Freitag B, Meister S, et al. Patient Acceptance of Prescribed and Fully Reimbursed mHealth Apps in Germany: An UTAUT2-based Online Survey Study. J Med Syst 2023; 47: 14.

26. Psychoula I, Singh D, Chen L, et al. Users’ Privacy Concerns in IoT Based Applications. In: 2018 IEEE SmartWorld, Ubiquitous Intelligence & Computing, Advanced & Trusted Computing, Scalable Computing & Communications, Cloud & Big Data Computing, Internet of People and Smart City Innovation (SmartWorld/SCALCOM/UIC/ATC/CBDCom/IOP/SCI). Guangzhou, China: IEEE, pp. 1887–1894.

27. Hair JF, Ringle CM, Sarstedt M. PLS-SEM: Indeed a Silver Bullet. Journal of Marketing Theory and Practice 2011; 19: 139–152.

28. Goffman E. Stigma: Notes on the Management of Spoiled Identity. Prentice-Hall, Inc., 1963.

29. Peer E, Brandimarte L, Samat S, et al. Beyond the Turk: Alternative platforms for crowdsourcing behavioral research. Journal of Experimental Social Psychology 2017; 70: 153–163.

30. Stablein T, Hall JL, Pervis C, et al. Negotiating stigma in health care: disclosure and the role of electronic health records. Health Sociology Review 2015; 24: 227–241.

31. Cheng Y-H, Yeh Y-J. Exploring radio frequency identification technology’s application in international distribution centers and adoption rate forecasting. Technological Forecasting and Social Change 2011; 78: 661–673.

32. Miltgen CL, Popovič A, Oliveira T. Determinants of end-user acceptance of biometrics: Integrating the “Big 3” of technology acceptance with privacy context. Decision Support Systems 2013; 56: 103–114.

33. Glozier N. Workplace Effects of the Stigmatization of Depression: Journal of Occupational & Environmental Medicine 1998; 40: 793–800.

34. Strømsvik N, Råheim M, Øyen N, et al. Stigmatization and Male Identity: Norwegian Males’ Experience after Identification as BRCA1/2 Mutation Carriers. Journal of Genetic Counseling 2010; 19: 360–370.

35. Bickford J, Barton SE, Mandalia S. Chronic genital herpes and disclosure…. The influence of stigma. Int J STD AIDS 2007; 18: 589–592.

36. Zacks S, Beavers K, Theodore D, et al. Social Stigmatization and Hepatitis C Virus Infection: Journal of Clinical Gastroenterology 2006; 40: 220–224.

37. Cuschieri S. The CONSORT statement. Saudi J Anaesth 2019; 13: 27.

38. Blanz M. Forschungsmethoden und Statistik für die Soziale Arbeit. Grundlagen und Anwendungen. 1. Aufl.

39. Hair JF, Hult GTM, Ringle CM, et al. Partial Least Squares Strukturgleichungsmodellierung: eine anwendungsorientierte Einführung.

40. Fornell C, Larcker DF. Evaluating Structural Equation Models with Unobservable Variables and Measurement Error. Journal of Marketing Reseach 1981; 18: 39–50.

41. Henseler J, Ringle CM, Sarstedt M. A new criterion for assessing discriminant validity in variance-based structural equation modeling. J of the Acad Mark Sci 2015; 43: 115–135.

42. Homburg C, Baumgartner H. Beurteilung von Kausalmodellen. Bestandsaufnahme und Anwendungsempfehlungen. Marketing ZFP 1995; 17: 162–176.

43. Sarstedt M, Henseler J, Ringle CM. Multigroup Analysis in Partial Least Squares (PLS) Path Modeling: Alternative Methods and Empirical Results. In: Sarstedt M, Schwaiger M, Taylor CR (eds) Advances in International Marketing. Emerald Group Publishing Limited, pp. 195–218.

44. Zhu M, Wu C, Huang S, et al. Privacy paradox in mHealth applications: An integrated elaboration likelihood model incorporating privacy calculus and privacy fatigue. Telematics and Informatics 2021; 61: 101601.

45. Li T, Slee T. The effects of information privacy concerns on digitizing personal health records: The Effects of Information Privacy Concerns on Digitizing Personal Health Records. J Assn Inf Sci Tec 2014; 65: 1541–1554.

46. Ancker JS, Witteman HO, Hafeez B, et al. The Invisible Work of Personal Health Information Management Among People With Multiple Chronic Conditions: Qualitative Interview Study Among Patients and Providers. J Med Internet Res 2015; 17: e137.

47. Heirman W, Walrave M, Ponnet K. Predicting adolescents’ disclosure of personal information in exchange for commercial incentives: an application of an extended theory of planned behavior. Cyberpsychol Behav Soc Netw 2013; 16: 81–87.

48. Brown S-A, Jouni H, Marroush TS, et al. Disclosing Genetic Risk for Coronary Heart Disease: Attitudes Toward Personal Information in Health Records. American Journal of Preventive Medicine 2017; 52: 499–506.

49. Shen N, Bernier T, Sequeira L, et al. Understanding the patient privacy perspective on health information exchange: A systematic review. International Journal of Medical Informatics 2019; 125: 1–12.

50. Hartmann M. Die Praxis des Vertrauens. 1. Aufl. Berlin: Suhrkamp, 2011.

51. Mcknight DH, Carter M, Thatcher JB, et al. Trust in a specific technology: An investigation of its components and measures. ACM Trans Manage Inf Syst 2011; 2: 1–25.

52. Weiss MG, Ramakrishna J, Somma D. Health-related stigma: Rethinking concepts and interventions. Psychology, Health & Medicine 2006; 11: 277–287.

53. Custers B, Dechesne F, Sears AM, et al. A comparison of data protection legislation and policies across the EU. Computer Law & Security Review 2018; 34: 234–243.

54. Bettiga D, Lamberti L, Lettieri E. Individuals’ adoption of smart technologies for preventive health care: a structural equation modeling approach. Health Care Manag Sci 2020; 23: 203–214.

